# Linking environmental antimicrobial resistance to human acquisition: evidence, knowledge gaps and a mechanistic framework

**DOI:** 10.64898/2026.10.13.26365397

**Authors:** Maria Alexa, Elise Hodbert, Jérémy Seurat, Marion Opatowski, Aleksandra Kovacevic, Lulla Opatowski, Laurence Armand-Lefèvre, Didier Hocquet, Elena Buelow

## Abstract

The contribution of environmental reservoirs to antimicrobial resistance (AMR) acquisition in humans remains poorly quantified, despite extensive evidence that resistant bacteria and resistance genes circulate through wastewater, surface waters, food and other environmental matrices. The central difficulty is no longer to demonstrate that these reservoirs are connected, but to quantify the direction and relative contribution of the pathways linking environmental AMR to human acquisition. Here, we synthesize the main processes that need to be represented at the environment–human interface and propose a mechanistic framework for extended-spectrum beta-lactamase-producing Escherichia coli (ESBL-Ec) linking human, water and food compartments.

The framework explicitly represents human-to-environment contamination and multiple environment-to-human exposure routes, including drinking water, recreational water exposure and irrigation-mediated food contamination. We apply the model to two contrasting illustrative settings, France and Madagascar, and use antibiotic-use, environmental-contamination and climate-related scenarios to explore how changes in specific drivers propagate across compartments. The simulations show strongly compartment-dependent responses, while sensitivity analyses identify wastewater treatment efficiency and interface-specific transmission, exposure and environmental-selection parameters as influential components of the system.

The framework is used to explore how different transmission pathways and environmental drivers can shape AMR dynamics across interconnected human and environmental compartments. By comparing contrasting scenarios and identifying the parameters that most strongly influence model behaviour, we highlight both potential leverage points and critical gaps in current empirical knowledge. More broadly, this work illustrates how decomposing One Health AMR into tractable reservoir interfaces can help structure future data collection and progressively improve our ability to quantify transmission between reservoirs.

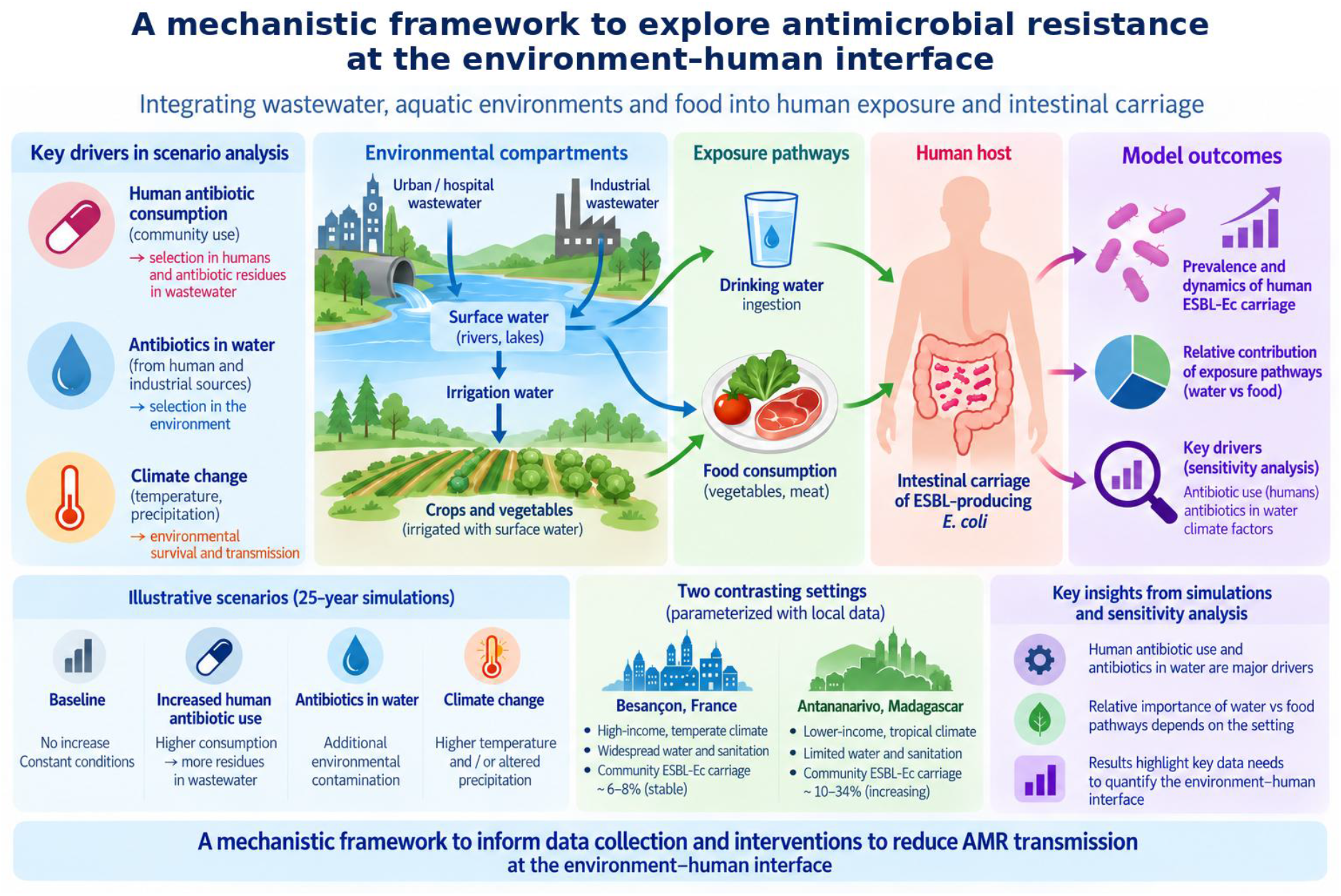

## 1. Introduction: why the environment–human interface remains difficult to quantify

Antimicrobial resistance (AMR) is increasingly recognized as an ecological phenomenon that extends far beyond clinical settings (Tacconelli et Pezzani 2019; Walsh 2018; Nadimpalli et al. 2018). Environmental microbial communities constitute vast reservoirs of antibiotic resistance genes (ARGs), where AMR can persist, diversify, and disseminate independently of direct human infection (Allen et al. 2010; Bengtsson-Palme et al. 2018). Rather than acting as passive sinks, anthropogenically impacted environments actively shape AMR dynamics through sustained selective pressures and frequent exchange of microorganisms and ARGs among interconnected microbial communities (Bengtsson-Palme et Larsson 2016; Catho et al. 2024; Thebo et al. 2017).

Extended-spectrum β-lactamase-producing *Escherichia coli* (ESBL-Ec) is an important resistant pathogen for humans globally (Naghavi et al. 2024). This resistant bacteria is prevalent in human and warm blooded animal microbiota and frequently detected in wastewater and aquatic environments (Blaak et al. 2014; Maric et al. 2025), making it a suitable indicator for studying how environmental conditions structure AMR persistence and dissemination.

To better understand the complex dynamics of AMR at the interface between environmental systems and human populations, modeling has provided an interesting framework. However, most modeling studies addressing AMR transmission across this interface tend to incorporate the environment within broader One Health frameworks that include human, animal, and environmental compartments (Booton et al. 2021; G. M. Knight et al. 2026; Lepper et al. 2022). Within such models the environmental dimension often remains relatively underexplored. When studies specifically address the environment–human interface, they typically focus on the impact of human activities on environmental contamination (Henriot et al. 2024) or on individual transmission pathways, such as exposure through drinking water or consumption of contaminated vegetables (O’Flaherty et al. 2018). While these approaches provide valuable insights into specific processes, their narrow scope limits a comprehensive understanding of AMR dynamics across interconnected ecosystems and their consequences for human health.

Against this background, we focus specifically on the environment–human interface as one tractable component of the wider One Health system. Our objective is threefold: (i) to synthesize the processes and exposure pathways that current evidence indicates should be represented at this interface; (ii) to formalize these processes in a mechanistic model linking human, water and food compartments; and (iii) to use illustrative simulations and sensitivity analysis to identify influential drivers, expose key assumptions and define the empirical data needed to quantify this interface. ESBL-Ec is used as a model organism because it is relevant to human carriage and disease and is repeatedly detected across wastewater, aquatic and food-associated environments. (Cavicchioli et al. 2019; Fernández Salgueiro et al. 2024; Kaba et al. 2020) (Levy et al. 2016; MacFadden et al. 2018)

## 2. From environmental reservoirs to human acquisition: current evidence and remaining gaps

The environment–human interface combines several distinct processes: selection and persistence of resistance in environmental microbial communities; release of human-associated bacteria and resistance determinants into environmental systems; transport and transformation across water and food pathways; and subsequent human exposure and acquisition. These processes are supported by different types of evidence and occur at different spatial and temporal scales. A useful mechanistic framework therefore needs to represent them separately before their combined contribution can be explored.

### 2.1 Environmental reservoirs, selection and persistence

Environmental pollution is a major driver of this process (Cavicchioli et al. 2019; Levy et al. 2016; Rodgers et al. 2019; Buelow et al. 2021). Antibiotics, biocides, metals, and other xenobiotic compounds released through urban, medical, industrial, and agricultural activities exert chronic and combinatorial selection of AMR on environmental microbiota. Such mixed exposure regimes favour the maintenance and mobilisation of ARGs, often at concentrations below inhibitory levels, and promote horizontal gene transfer (HGT) within complex microbial communities (Andersson et al. 2020; Andersson et Hughes 2014; Jutkina et al. 2018; Rodgers et al. 2019). Hence, selection of AMR in the environment is not limited to antibiotics alone, but is strongly influenced by co-selective and cross-selective pressures exerted by non-antibiotic pollutants (Cao et al. 2020; Cheng et al. 2019; Dickinson et al. 2019).

Among environmental compartments, water and wastewater systems occupy a central position in AMR dissemination (Buelow et al. 2020; Larsson et Flach 2022; Buelow et al. 2021). Wastewater integrates inputs from human populations, hospitals, industry, and agriculture, concentrating ARGs, antibiotic-resistant bacteria (ARB), and selective agents within highly connected microbial ecosystems(Flandroy et al. 2018; McInnes et al. 2021; Manaia et al. 2018). Wastewater treatment plants and receiving waters therefore function as interfaces between human-associated resistomes and environmental reservoirs, enabling persistence, reshuffling, and downstream transmission of resistance determinants.

Together, these observations justify representing wastewater and connected aquatic environments not simply as passive recipients of human-derived AMR, but as compartments in which resistant and susceptible populations can persist, be exposed to selective pressures and contribute to downstream exposure pathways.

### 2.2 Bidirectional exchange between humans and environmental compartments

The human-environment interface is intrinsically bidirectional. Human carriage contributes resistant bacteria to wastewater through faecal shedding, while environmental contamination can return to human populations through drinking water, recreational contact with surface waters and contaminated food. These routes differ in exposure frequency, dose and probability of acquisition, and therefore should not be collapsed into a single generic environmental transmission term.

Food provides an additional indirect route. Irrigation with contaminated surface water or treated wastewater can transfer resistant bacteria to crops, while animal-derived food can contribute independently to human exposure. In the present framework, these pathways are represented explicitly where data permit, allowing their contribution to be examined separately rather than inferred from a single environmental reservoir.

### 2.3 Context, climate-related perturbations and data limitations

The impact of these pathways is expected to depend strongly on context. Wastewater treatment, sanitation infrastructure, antibiotic consumption, environmental contamination, water use and human behaviour differ between settings and can modify both environmental AMR dynamics and opportunities for human exposure. Climate-related perturbations can act through several of these mechanisms simultaneously: warming can modify bacterial growth and persistence, drought can alter water use and recreational exposure, water scarcity can increase wastewater reuse, and extreme precipitation can reduce effective wastewater treatment through overflow or bypass events.

Previous studies emphasized that reducing bacterial transmission throughout the One Health compartments (humans, animals and the environment) is essential to lowering AMR prevalence in humans. Yet our understanding of the One Health transmission dynamics and the quantitative contribution of the different compartments remains limited by the lack of longitudinal data monitoring jointly AMR prevalence in humans, but also antibiotic use, and environmental exposure across multiple sectors and settings (G. Knight et al. 2025; Berendonk et al. 2015a; Rousham et al. 2018). This highlights the need for approaches that integrate environmental exposure, microbial ecology, and human health data to capture the complex pathways linking the different environmental reservoirs and clinical AMR.

The resulting evidence base is therefore asymmetric: the existence of many individual pathways is well supported, but quantitative estimates of their direction, strength and interaction are sparse. Comparable longitudinal measurements across human and environmental compartments are particularly limited. This is the specific gap addressed by the model below: the framework is intended to formalize plausible pathways, explore their consequences under explicit assumptions and identify which missing measurements are most important, rather than to provide a calibrated forecast of future AMR prevalence.

## 3. Proposed mechanistic framework for the environment–human interface

We developed a mathematical model to formalize persistence, selection and transmission of ESBL-Ec across the environment–human interface. The framework was deliberately constructed as an interface-specific component of a broader One Health modelling strategy: it does not attempt to represent all human, animal and environmental reservoirs simultaneously. Instead, it captures the principal pathways identified in the evidence synthesis above and provides a tractable structure in which their relative influence, interactions and data requirements can be explored.

### 3.1. Model overview

The model provides a first-order representation of ESBL-Ec dynamics across interconnected human, water and food compartments. It explicitly represents bacterial exchange between compartments while allowing demographic, environmental and behavioural drivers to modify transmission and persistence.

Three sub-models. The framework links water, vegetables and the general human population, with contaminated meat represented as an external exposure input rather than a dynamically modelled food reservoir. Water compartments are described by ESBL-Ec concentrations (CFU/100 mL), whereas vegetable contamination and human carriage are represented as prevalences. The linked submodels are described below and illustrated in Figure 1.

**Figure 1.**
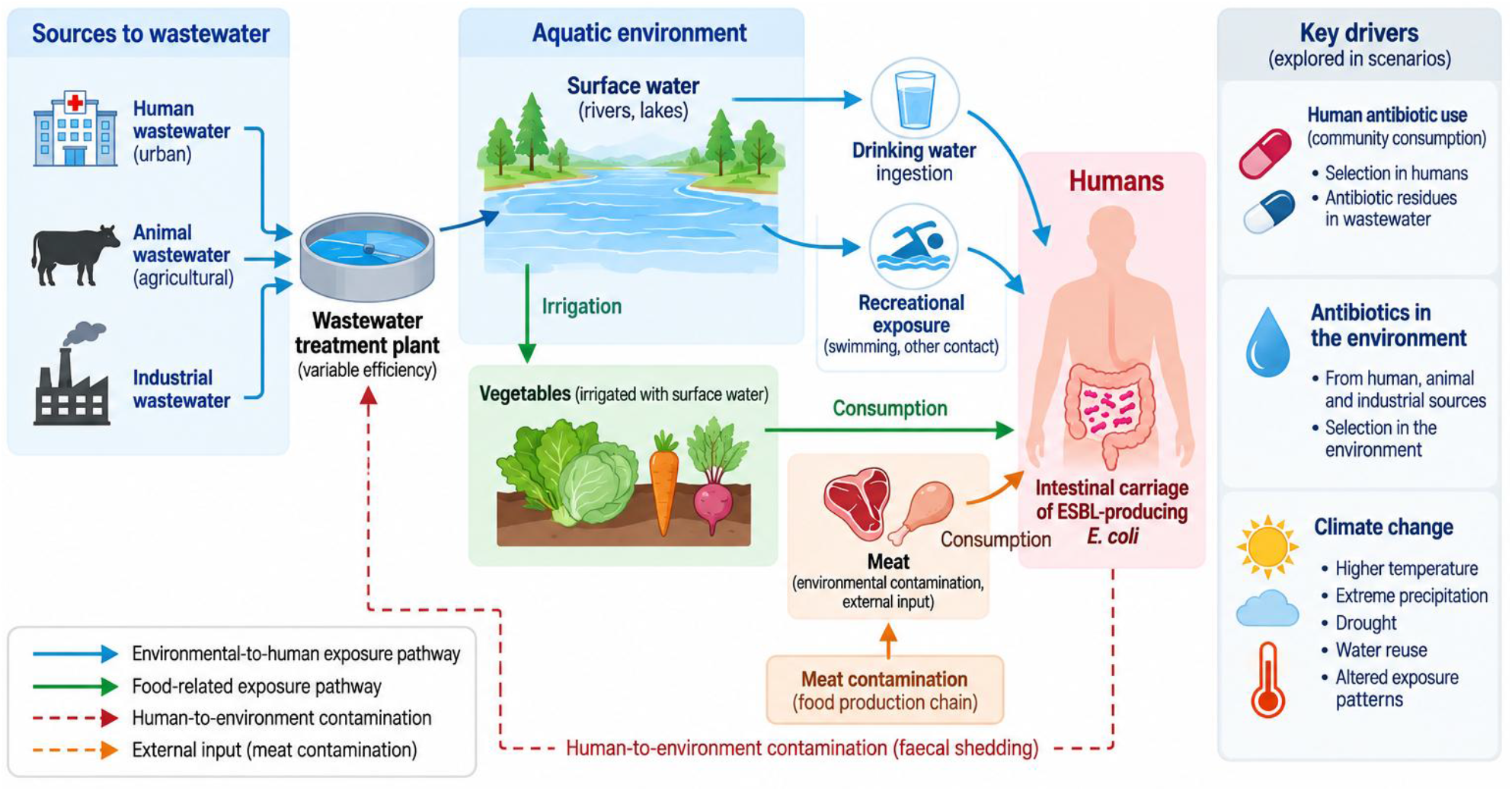
Simplified overview of the environment–human interface model. Urban/hospital, animal and industrial wastewater contribute to surface-water contamination following treatment. Environmental exposure occurs through drinking and recreational water and irrigated vegetables; meat contamination is represented as an independent external exposure input. Human faecal shedding contributes to urban/hospital wastewater. Antibiotic use and climate-related factors modify model processes.

Two contrasting settings. As AMR spread is highly heterogeneous across countries and is largely driven by differences in infrastructure, sanitation and socioeconomic conditions (Collignon et al. 2018; Rahbe et al. 2023), we used the framework to explore ESBL-Ec dynamics in two contrasting settings: a high-income setting (the 120,000-inhabitant city of Besançon, France) and a low-income setting (the 1,600,000-inhabitant city of Antananarivo, Madagascar). For both settings, antibiotic-related and climate-related scenarios were explored over a 25-year simulation horizon; these applications are illustrative rather than forecasts. (Collignon et al. 2018; Rahbe et al. 2023)

#### 3.1.1. General human population model

The general human population submodel is illustrated in Figure 2. It incorporates the key variables influencing the transmission of ESBL-Ec and subsequent carriage by humans, accounting for both human-to-human and environment-to-human exposure pathways.

**Figure 2.**
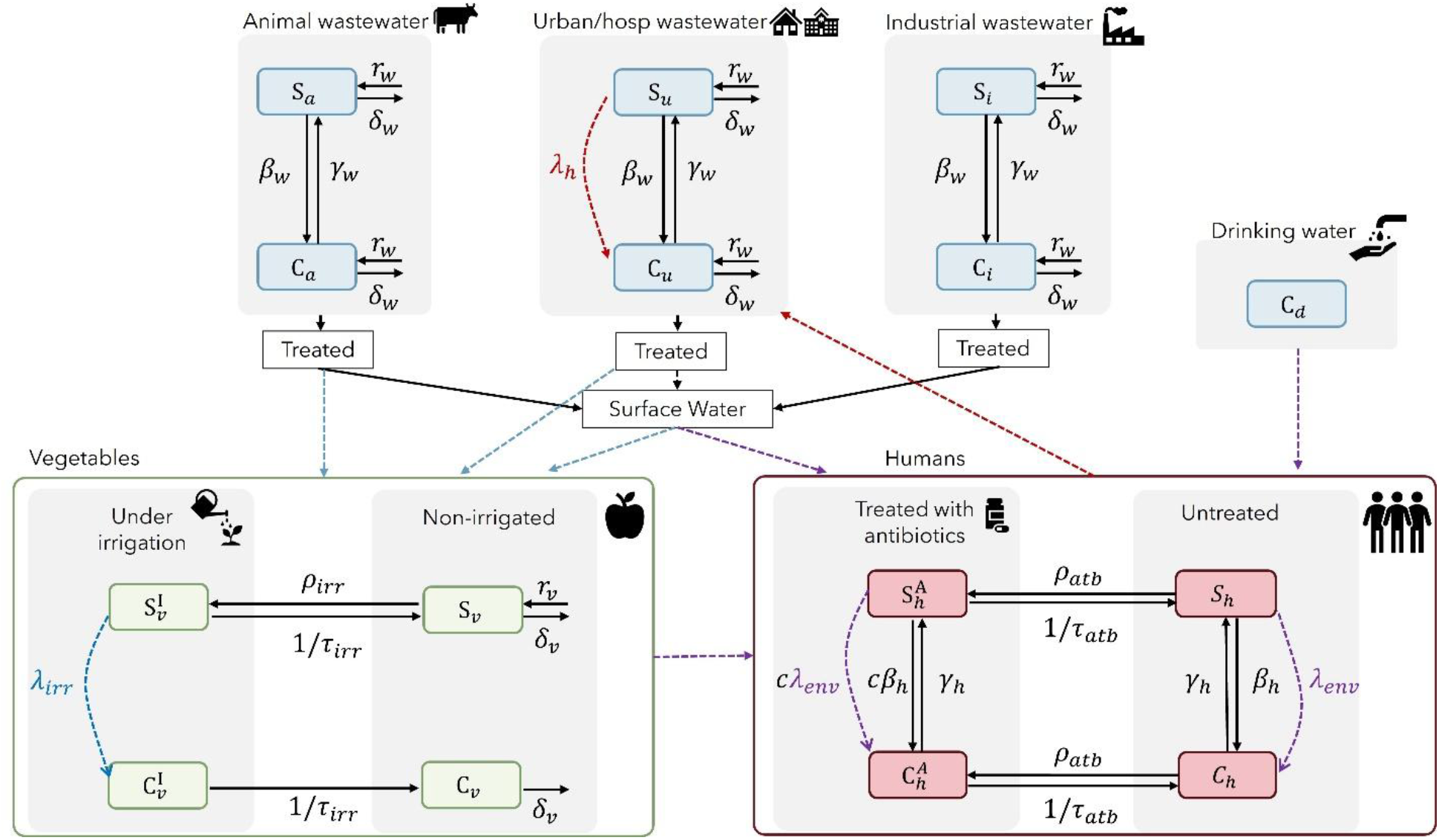
Conceptual model of the most significant transmission routes of ESBL-producing E. coli between water, food and human. Dashed lines represent contributions of reservoir-to-reservoir contamination. Purple dashed line represents environment-to-humans contamination, red dashed line represents human-to-wastewater contamination and blue dashed line represents wastewater-to-vegetables contamination.

Individuals were first classified according to colonization status as either carriers (*C*_ℎ_) or non-carriers (*S*_ℎ_) of ESBL-Ec. As *E. coli* is a commensal intestinal bacterium found in 90% of humans (Tenaillon et al. 2010), we assume that all non-carriers are colonized with antibiotic-sensitive *E. coli*, as in previous studies (Rahbe et al. 2025). Non-carriers may acquire ESBL-Ec through inter-individual transmission (i.e, from a human carrier) at a daily per-capita rate *β*_ℎ_ or through contact with environmental sources at rate *λ_env_*. Carriers may clear colonization and return to the non-carrier state at a constant rate *γ*_ℎ_. The population is further stratified according to antibiotic exposure, distinguishing individuals receiving antibiotic treatment (*S*_ℎ_*^A^*, *C*_ℎ_*^A^*) from those not receiving treatment (*S*_ℎ_, *C*_ℎ_). We consider the proportion of individuals *ρ_atb_* who initiate antibiotic therapy each day, with treatment lasting for a fixed duration of *τ_atb_* days. During treatment, antibiotic exposure induces microbiome dysbiosis (de Nies et al. 2023), which increases the risk of ESBL-Ec acquisition by a factor *c.* Besides, the modelling of exposure of humans to environmental sources of resistance takes into account bathing, drinking, consumption of plants and animal products (Supplementary Table 1).

#### 3.1.2. Water submodel concerning non surface waters

Three wastewater sources are considered: urban/hospital (*u*), animal (*a*), and industrial (*i*). For each source *w* ∈ {*u*, *a*, *i*}, we developed a deterministic model to describe the evolution of concentrations of antimicrobial-susceptible (*S_w_*; non-ESBL) and antimicrobial-resistant (*C_w_* ESBL-producing) *E. coli* in water.

The framework builds on previous models describing within-host bacterial population dynamics (Blanquart 2019a). Similar bacterial population-dynamic approaches have also been applied to stored agriculture waste (Baker et al. 2016) and subsequent adaptations to wastewater (Sutradhar et al. 2021).

##### Bacterial growth

Each water compartment (*w*) is divided into two bacterial populations: Ec in water *S_w_* and ESBL-Ec in water *C_w_*. Both populations are assumed to grow logistically with a shared carrying capacity and the same intrinsic growth rate, assuming no fitness difference between susceptible and resistant bacteria. Resistance spreads via HGT *β_w_* from resistant to susceptible bacteria through plasmid conjugation. Antimicrobial resistance genes can be lost at rate *γ_w_*. Both susceptible and resistant bacteria decay at rate *δ_w_*. Antibiotic exposure modifies bacterial growth through the antibiotic-dependent factors, which represent the relative growth capacity of susceptible and resistant bacteria under fluoroquinolone exposure. The human gut microbiome contributes bacteria to wastewater through fecal shedding.

##### Antibiotic effects on bacterial population dynamics

Antibiotic effects were modelled as an antibiotic-dependent increase in bacterial removal using a Hill function based on antibiotic concentrations in water. Ciprofloxacin concentrations measured in wastewater and heavily polluted receiving waters have been reported to select for fluoroquinolone-resistant bacteria (Haenni et al. 2022; Hayes et al. 2022). Because ESBL-Ec are frequently co-resistant to fluoroquinolones (Critchley et al. 2020), we used ESBL-Ec. as a model organism to describe the relationship between environmental fluoroquinolone concentrations and bacterial growth dynamics.

The antibiotic effects on susceptible and resistant bacteria were represented by growth reduction factors. These factors modify bacterial growth rates by accounting for the reduction in growth caused by antibiotic exposure.

##### Human microbiome contribution to urban and hospital wastewater

Human shedding contributes through feces to the contamination of the urban and hospital (*u*) wastewater compartment with ESBL-Ec. The human shedding affects the urban/hospital (*u*) wastewater compartment but does not directly contribute to animal wastewater (*a*), industrial wastewater (*i*) in the present model.

#### 3.1.3. Wastewater treatment, mixing, and bacterial decay in surface waters

The concentration of ESBL-Ec in surface water at the potential human exposure site, denoted (*C_s_*), was estimated by modelling the fate of ESBL-Ec from wastewater discharge to the receiving water body. The modelling framework consists of three successive steps: (i) wastewater treatment, (ii) mixing of treated effluents in the receiving water body, and (iii) bacterial decay during transport in the environment, arriving at the concentration at the potential human exposure site, denoted (*C_s_*). Three wastewater sources are considered: urban/hospital (*u*), animal (*a*), and industrial (*i*). For each source *w* ∈ {*u*, *a*, *i*}, the untreated wastewater contains an ESBL-Ec concentration (*C_w_*).

#### 3.1.4. Vegetable submodel

The vegetable submodel describes the contamination of vegetables with ESBL-Ec,, following irrigation with contaminated water, accounting for the exposure to ESBL-Ec by irrigation using surface water, urban treated water, and animal wastewater.

Vegetables are first classified according to their irrigation status, as either non-irrigated (*S_v_*) or irrigated (*S_v_^I^*). Non-irrigated vegetables enter the irrigated state at a rate *ρ_irr_*. During irrigation, vegetables may be exposed to contaminated water and become carriers of ESBL-Ec at rate *λ_irr_*. Once the irrigation period ends, exposed vegetables transition to the contaminated compartment *C_v_* at a fixed rate *τ_irr_*, while unexposed vegetables return to the non-irrigated compartment (*S_v_*). Both contaminated (*C_v_*) and non-contaminated (*S_v_*) vegetables are harvested at a rate *γ_v_* and new uncontaminated vegetables are continuously introduced into the system at a rate *r_v_*.

### 3.2. Application to two contrasting socio-economic settings

We applied the framework to two contrasting socio-economic and environmental settings for which setting-specific data were available: Besançon, France (population 0.12 million), representing a high-income temperate setting with widespread access to drinking water and wastewater treatment, and Antananarivo, Madagascar (population 1.6 million), representing a lower-income tropical setting with more limited access to drinking water and sanitation infrastructure.

#### Human model parameterization

In Europe, community ESBL-Ec carriage increased during the early 2000s (Woerther et al. 2013; Bezabih et al. 2021) before stabilizing at approximately 6–8% from around 2010 onwards (Blanquart 2019b; Bezabih et al. 2021; Castanheira et al. 2021; Pennings 2025). The human transmission rate for the French setting was therefore calibrated to maintain community ESBL-Ec prevalence within this range.

In Madagascar, community ESBL-Ec carriage is substantially higher. Prevalence was estimated at 10.1% in a community survey in 2011 (Herindrainy et al. 2011and at 34% among healthy pregnant women in 2018–2019 (Milenkov et al. 2021). Although these estimates are not directly comparable because of differences in the study populations, they are consistent with the broader pattern reported across Africa, where community carriage has been estimated at approximately 20% and has continued to increase (Woerther et al. 2013; Bezabih et al. 2021). The human transmission rate for the Madagascar setting was therefore calibrated to reproduce an increasing prevalence trajectory.

### 3.3. Scenario exploration

The model was simulated over a 25-year period under alternative scenarios designed to explore how changes in human antibiotic use, environmental antibiotic contamination and climate-related drivers affect ESBL-Ec dynamics across compartments.

A baseline scenario (B0) provides the reference against which alternative scenarios are compared. It assumes limited inter-reservoir interactions, constant human antibiotic use, no additional antibiotic residues in water and no climate-related perturbation. Baseline parameter values for both settings are provided in Supplementary Tables S1–S3 and initial conditions in Supplementary Table S4.

#### 3.3.1. Antibiotic use and environmental contamination scenarios

We investigated the impact of antibiotic changes in consumption and environmental antibiotic contamination through three scenarios representing different patterns of selective pressure in the human and environmental compartments.

Scenario A1 considers temporal changes in human antibiotic consumption, to assess the impact of increasing consumption patterns, while assuming the absence of antibiotics in the aquatic environment.

Scenario A2 assumes constant antibiotic exposure in both humans and the environment, with fixed human antibiotic consumption and a constant fluoroquinolone concentration in water, representing persistent environmental contamination.

Scenario A3 links temporal changes in human antibiotic consumption to environmental contamination, such that both human antibiotic consumption and environmental fluoroquinolone concentrations vary over time. This scenario represents a coupled human–environment system in which antibiotic use directly influences environmental contamination.

The three scenarios are summarized in Table 1.

**Table 1.** Antibiotic use and environmental contamination scenarios evaluated in the model. Climate change scenarios and associated model assumptions. The model was implemented using the R package odin.

| Scenario | Name | Description and parameterization |
| --- | --- | --- |
| **Baseline** |  |  |
| **B0** | Baseline | Limited inter-reservoir interactions; constant human antibiotic use ($\rho_{atb}$); no antibiotics in water; no climate perturbation |
| **Antibiotic use scenarios** |  |  |
| **A1** | Variable human antibiotic use | Human antibiotic use varies:<br>$\rho_{atb}(t)$ increases linearly from its baseline value to twice the baseline value over the simulation period; |
| **A2** | Persistent environmental contamination | Constant human antibiotic use:<br>$\rho_{atb}(A2) = \rho_{atb}(B0)$<br>and water contamination:<br>- In France $FQ_u(A2) = 120$ ng/L<br>- In Madagascar $FQ_u(A2) = 220$ ng/L |
| **A3** | Coupled human–environment dynamics | Human antibiotic use varies (A1) and water contamination vary together. Water contamination has the same linear trend as the human antibiotic consumption. |
| **Climate change scenarios** |  |  |
| **C1** | Warming | Increase of water temperature:<br>- Increase of HGT in water: $\beta_w(C1) = 1.05 \times \beta_w$<br>- Increase of bacterial growth rate in water: $r_w(C1) = 1.05 \times r_w$ ;<br>- Increase of carrying capacity in water: $N_{max}(C1) = 1.05 \times N_{max}$ |
| **C2** | Warming<br>+ Water reuse | C1 + increased irrigation with treated wastewater:<br>$\beta_{irr}^{tr}(C2) = 1.25 \times \beta_{irr}^{tr}$ |
| **C3** | Warming<br>+ Extreme precipitation | C1 + Reduced wastewater treatment efficiency due to overflow/bypass:<br>- In France: $effTP(C3) = 0.8$<br>- In Madagascar: $effTP(C3) = 0$ |
| **C4** | Warming<br>+ Increased drought periods | C1 + increased human contacts with surface water:<br>- $\beta_{swim}(C4) = 1.25 \times \beta_{swim}$<br>- $\beta_{shed}(C4) = 1.25 \times \beta_{shed}$ |
| **C5** | Warming<br>+ Water reuse<br>+ Extreme precipitation<br>+ Drought exposure | C1+C2+C3+C4 |

#### 3.3.2. Climate change scenarios

For both settings, we simulated the potential impact of climate change up to 2050 (25 years) by defining scenarios incorporating increases in temperature and associated environmental changes affecting transmission dynamics, including extreme precipitation, drought, and shifts in water use practices. These are detailed below and in Table 1.

The first scenario considers only an increase in water temperature (C1 – Warming) (van Vliet et al. 2013; Liu et al. 2020), and reflect its putative influence on microbial dynamics in aquatic environments. Elevated temperatures are associated with increased bacterial growth rates, enhanced HGT and greater environmental carrying capacity, as reported in an environmental microbiology and antimicrobial resistance study (Kalanxhi et Laxminarayan 2026).

The warming with water reuse scenario (C2 – Warming + water reuse) reflects increased reliance on treated wastewater for irrigation under drought conditions (Álvarez-Fraga et al. 2026). While current reuse remains limited in France, policy targets anticipate increased adoption in the coming years (Mallard et al. 2025). Increased reuse may enhance environmental dissemination of antibiotic residues and resistant bacteria in agricultural systems through irrigation practices.

The warming with extreme precipitation scenario (C3 – Warming + extreme precipitation) assumes an increase in both the frequency and intensity of heavy rainfall events (Intergovernmental Panel on Climate Change (IPCC) 2023). Extreme rainfall leads to combined sewer overflows and bypass events, reducing treatment efficiency and increasing the discharge of untreated or partially treated wastewater into surface waters (Giakoumis et Voulvoulis 2026). Such effects have been documented during storm events, including sharp increases in fecal and microbial contamination in river systems (Passerat et al. 2011).

Finally, warming coupled with increased periods of drought scenario (C4 – Warming + increased periods of drought) assumes increase of human exposure to surface waters. This assumption is supported by the increasing recreational use of urban rivers during hot summer periods and heatwaves, as river bathing is increasingly promoted as part of strategies to improve urban livability and adaptation to climate change.

The combined scenario (C5 – Warming + water reuse + extreme precipitation + drought exposure) represents the cumulative effects of climate change on transmission dynamics. It combines increased environmental dissemination through wastewater reuse and storm-related overflows with greater human exposure to surface waters during droughts and heatwaves.

Together, these scenarios represent key pathways through which climate change may influence transmission dynamics across human and environmental reservoirs.

### 3.4. Sensitivity analysis

A sensitivity analysis was performed to identify the parameters with the greatest influence on model predictions. Partial rank correlation coefficients (PRCCs) were used to quantify the relationship between model parameters and model outputs. All parameters were sampled simultaneously from normal distributions centered on their baseline values, with standard deviations corresponding to 10% of each baseline values. A total of 1,000 parameter sets were generated, and the model was simulated over a 25-year period for each parameter set. PRCCs were then computed between parameter values and the predicted ESBL-Ec prevalence in humans, vegetables and urban wastewater, with confidence intervals estimated using 100 bootstrap replicates.

## 4. Illustrative use of the framework: scenario exploration and sensitivity analysis

Scenario outcomes

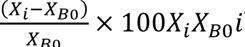 The model was simulated over a 25-year horizon under each scenario. These simulations are used to compare model behaviour under alternative assumptions rather than to forecast ESBL-Ec prevalence or concentration in France or Madagascar at a specific future date. Scenario outcomes were examined across the full simulated dynamics and relative to the baseline scenario (B0); detailed endpoint comparisons are reported in Supplementary Tables S5-S8.

### 4.1 Antibiotic use and environmental contamination reveal compartment-specific responses

The antibiotic-related scenarios illustrate how selective pressure introduced in different parts of the system can propagate across compartments (Figure 3). Increasing human antibiotic consumption primarily changed predicted human carriage, whereas environmental antibiotic contamination produced a stronger response in the wastewater compartment. The coupled scenario generated the largest changes across compartments in both settings (Supplementary Tables S5-S6). These results should be interpreted as consequences of the model structure and parameter assumptions rather than as quantitative predictions; their value lies in showing how human and environmental selective pressures can generate different system-level responses.

**Figure 3.**
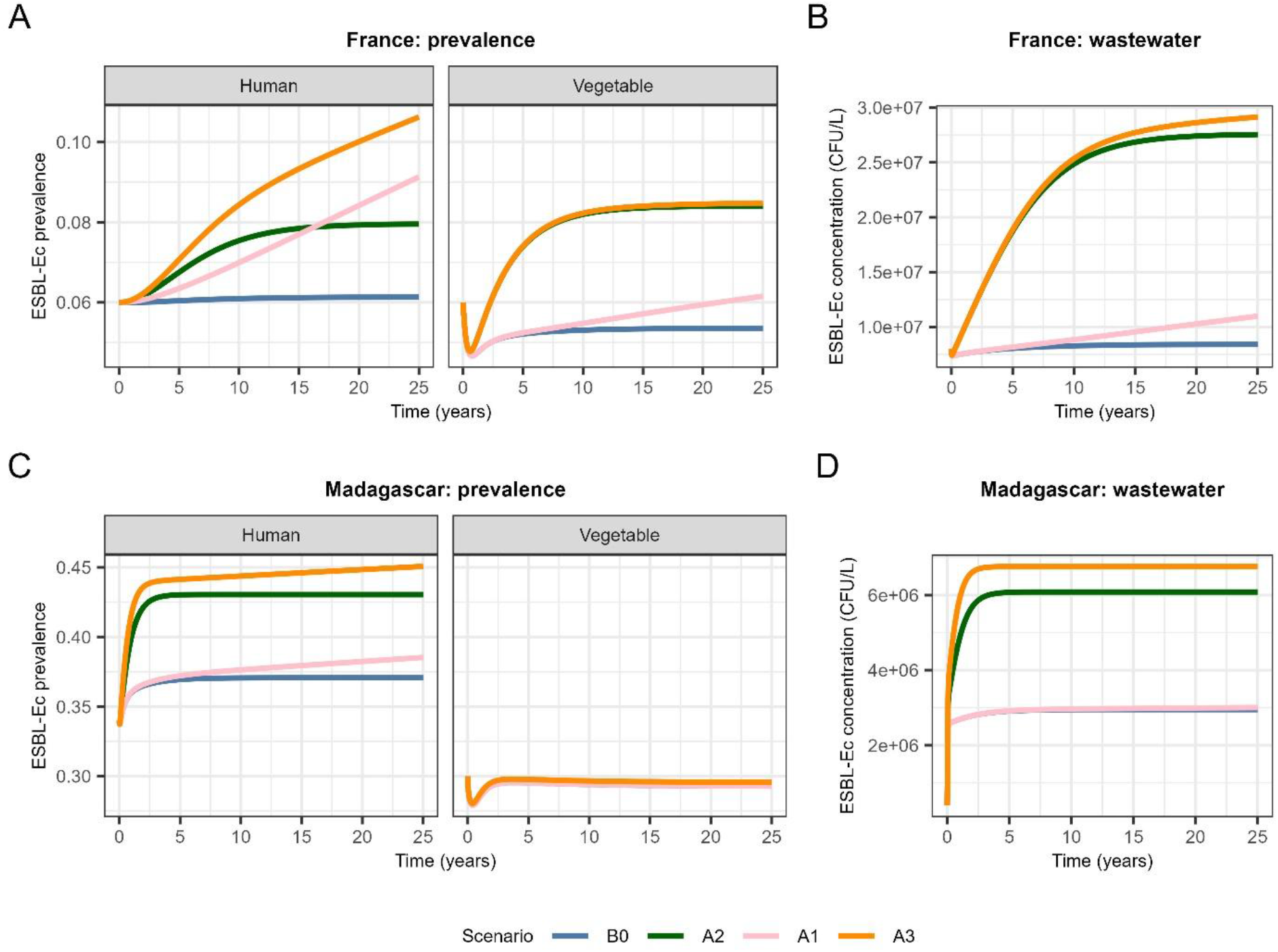
Effects of human antibiotic use and environmental antibiotic contamination on ESBL-E. coli prevalence and concentration. Model predictions over 25 years under baseline and antibiotic-related scenarios. Panels show predicted ESBL-Ec prevalence in humans and vegetables in (A) France and (C) Madagascar, and predicted ESBL-E. coli concentrations in urban wastewater in (B) France and (D) Madagascar. Scenarios were defined as follows: B0, baseline conditions, with constant human antibiotic use and no additional antibiotic contamination in wastewater; A1, variable human antibiotic use but no antibiotic contamination in wastewater; A2, environmental contamination, with constant human antibiotic use; and A3, coupled human– environment dynamics.

### 4.2 Climate-related perturbations act through multiple pathways

Climate-related scenarios produced smaller changes under warming alone than when warming was combined with changes in wastewater treatment, water reuse or recreational exposure (Figure 4; Supplementary Tables S7-S8). This illustrates an important feature of the framework: climate-related effects on AMR need not arise from temperature alone, but can emerge through simultaneous changes in infrastructure, environmental contamination and human behaviour. The magnitude of these responses remains conditional on the scenario assumptions and should therefore be interpreted comparatively rather than as a forecast.

**Figure 4.**
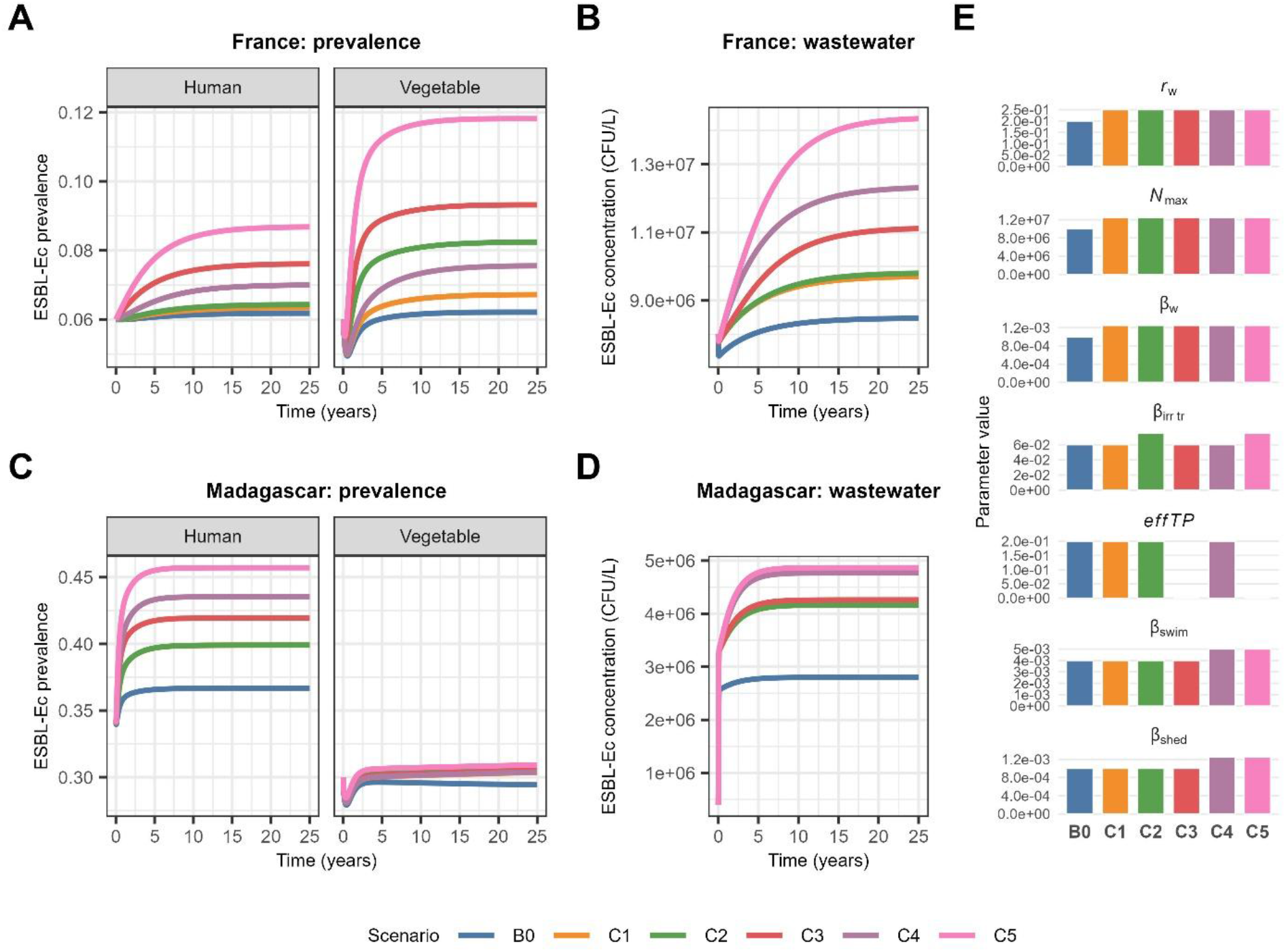
Effects of climate change scenarios on ESBL-Ec prevalence and concentration. Model predictions from 2025 to 2050 under baseline and climate change scenarios. Panels show predicted ESBL-EC prevalence in humans and vegetables in (A) France and (C) Madagascar, and predicted ESBL-Ec concentrations in (B) France and (D) Madagascar. Panel (E) illustrates the values of the parameters used for the different scenarios in Madagascar.The parameters shown are the bacterial growth rate in water, the carrying capacity, the gene transfer rate in wastewater, the transmission rate from treated water to vegetables, the efficacy of the WWTP, the transmission rate from surface water to humans, the transmission rate from humans to water. Scenarios were defined as follows: B0, baseline conditions (no additional climate-related perturbation); C1, warming alone; C2, warming combined with increased water reuse; C3, warming combined with increased extreme precipitation; C4, warming combined with increased drought-related exposure; and C5, warming combined with increased water reuse, extreme precipitation, and drought-related exposure.

### 4.3 Sensitivity analysis identifies influential parameters and data priorities

The sensitivity analysis identified distinct influential parameters across compartments (Figure 5). Human prevalence was primarily associated with parameters describing human-to-human transmission and natural clearance, whereas vegetable prevalence was strongly influenced by wastewater treatment efficiency and irrigation-related parameters. Predicted ESBL-Ec concentrations in wastewater were particularly sensitive to HGT, fluoroquinolone exposure, bacterial removal and wastewater treatment efficiency. Wastewater treatment efficiency was one of the few parameters associated with outcomes across several compartments, illustrating how sensitivity analysis can identify candidate leverage points as well as parameters for which improved empirical estimates would be especially informative.

**Figure 5.**
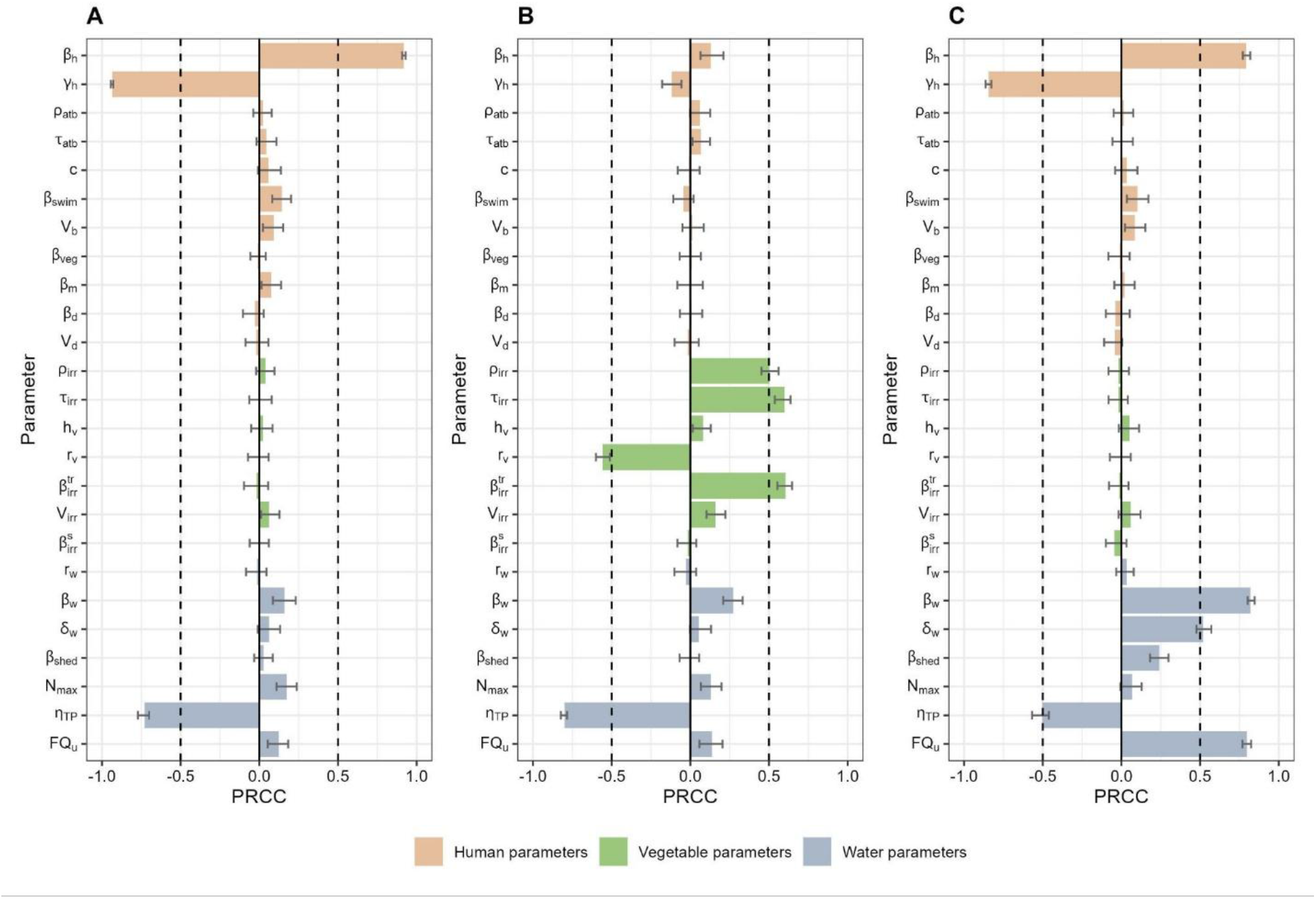
Sensitivity analysis for ESBL-Ec prevalence after 25 years. Partial rank correlation coefficients (PRCCs) between model parameters and predicted ESBL-Ec prevalence after 25 years for (A) humans, (B) vegetables and (C) urban WW. Parameters were sampled simultaneously from normal distributions centred on their baseline values (SD = 10%). Bars represent PRCC values and error bars indicate 95% confidence intervals estimated from 100 bootstrap samples. Parameters are colour-coded according to the compartment they describe (human, orange; vegetables, green; water, blue).

## 5. Discussion: what an interface-specific model adds

The central contribution of this work is not a forecast of future ESBL-Ec prevalence, but an explicit formalization of the environment–human interface. We first considered the evidence supporting environmental selection, human-to-environment release and multiple environment-to-human exposure pathways, and then translated these processes into a tractable mechanistic framework. This follows the broader logic of decomposing One Health AMR into defined reservoir interfaces that can be studied, parameterized and challenged individually before being connected within larger multi-reservoir systems. (Booton et al. 2021; O’Flaherty et al. 2018; Henriot et al. 2024)

The simulations show why this decomposition is useful. Drivers introduced in one compartment do not affect all outcomes equally: human transmission parameters primarily shape human carriage, irrigation-related processes influence food contamination, and environmental selection and HGT parameters are most visible in wastewater dynamics. At the same time, cross-compartment parameters such as wastewater treatment can influence several parts of the system. The framework therefore provides a way to ask where an observed change is expected to propagate, which pathways mediate that propagation and which measurements are required to test the underlying hypothesis.

The antibiotic scenarios illustrate that selective pressures acting in humans and wastewater can generate different responses across connected compartments. Their combined effects underline the importance of representing human antibiotic use and environmental contamination separately. These simulated differences are conditional on the specified model assumptions and should not be interpreted as direct evidence of environmental acquisition or as calibrated intervention effects.

In our simulations, investigated rising temperature scenarios alone had comparatively modest effects on predicted ESBL-Ec prevalence in both settings. In contrast, scenarios combining rising temperatures with additional environmental perturbations, including drought, flooding, water reuse, and increased recreational water exposure, consistently produced larger increases of ESBL-Ec concentrations across environmental compartments (Figure 4). These results suggest that climate change is unlikely to influence AMR transmission through a single mechanism but rather by modifying multiple environmental exposure pathways simultaneously. Increasing temperatures may enhance environmental bacterial survival and growth and antibiotic concentration, while extreme weather events can alter wastewater infrastructure performance, increase sewer overflows, and promote the dissemination of resistant bacteria into receiving waters (Singer et al. 2016; Environment 2024; Berendonk et al. 2015b). Likewise, increasing recreational use of surface waters during periods of elevated temperatures may increase opportunities for human exposure to environmental AMR (Leonard et al. 2022). This underscores the need for future global AMR-related risk assessments to account for climate change–driven shifts in human behavior.

Sensitivity analysis further distinguished the parameters most influential for each compartment: human transmission and clearance for human carriage, irrigation and wastewater treatment for vegetable contamination, and horizontal gene transfer, fluoroquinolone exposure and bacterial removal for wastewater concentrations. Wastewater treatment efficiency influenced several model outcomes, making it a priority for improved empirical parameterization rather than a demonstrated intervention effect.

An important strength of this study lies in the explicit integration of multiple environmental exposure pathways within a One Health framework. By combining environmental contamination, human behavior, food contamination, and climate-related changes within a single modelling framework, the model enables direct comparison of the relative contribution of different transmission routes and intervention strategies. Its modular structure also facilitates future refinement as new epidemiological or environmental data become available, allowing additional reservoirs, exposure pathways, or mitigation strategies to be incorporated without fundamentally altering the underlying framework.

A major result of this formalization is the visibility it gives to current data gaps. Several interface parameters cannot yet be estimated robustly from directly comparable studies. Environment-to-human acquisition probabilities, human shedding into wastewater, persistence between discharge and exposure sites, contamination during irrigation and the effect of environmental selective concentrations are rarely measured together. Consequently, several model parameters necessarily rely on heterogeneous literature, setting-specific assumptions or indirect estimates. These limitations should be considered when interpreting scenario magnitude, but they also define a concrete empirical research agenda. (Haverkate et al. 2017) (Schets et al. 2011; Spencer et al. 2022)

Future studies should therefore prioritize matched and longitudinal measurements across the interface: human carriage together with wastewater and receiving-water contamination; exposure frequency and dose for drinking and recreational water; contamination and persistence along wastewater-reuse and food pathways; environmental concentrations of selective agents; and wastewater treatment performance. Sensitivity analysis can help prioritize these measurements by identifying the uncertain parameters to which outcomes are most responsive and can support study design by indicating relevant compartments and time scales for sampling.

Several additional limitations should be considered. The framework is intentionally simplified and does not explicitly represent microbial community interactions, strain-level evolutionary dynamics or all possible environmental reservoirs. Parameterization is heterogeneous in quality and is substantially better supported for some processes than for others. The two settings are therefore used to illustrate how the same mechanistic structure behaves under contrasting epidemiological, infrastructural and environmental assumptions, not as validated city-level forecasting models. Extension to other settings will require locally relevant epidemiological, environmental and behavioural data and, where appropriate, additional interface-specific compartments.

## 6. Conclusion

The environment-human contribution to AMR cannot be understood from the presence of resistance determinants in individual reservoirs alone. It requires explicit representation of the pathways connecting human carriage, wastewater, aquatic environments, food contamination and human exposure. By combining a narrative synthesis of these mechanisms with an interface-specific mechanistic model, we provide a framework for making these hypotheses explicit and testable. The simulations illustrate how different drivers can propagate through different compartments, while sensitivity analysis identifies parameters for which improved empirical evidence would most strengthen inference. This framework is therefore intended as a platform for study design, progressive parameter refinement and evaluation of intervention hypotheses at the environment–human interface, and as one component of a broader strategy to build tractable One Health AMR models through connected reservoir interfaces (*WHO Bacterial Priority Pathogens List 2024* 2024).

## Data Availability

All model inputs and simulation results reported in this study are provided in the manuscript and supplementary material. Sources of publicly available parameter values are cited therein. No individual participant data were collected or used.

## Acknowledgements

The authors acknowledge the PROMISE network for facilitating the scientific exchanges and collaborative discussions during the PROMISE hackathon that led to the development of this work.

