## Supplementary Data for "Linking environmental antimicrobial resistance to human acquisition: evidence, knowledge gaps and a mechanistic framework"

Appendix 1

### Table S1. Characteristics of parameters used for the general population model

| **Symbol** | **Parameter** | **Unit** | **Value** | | **References** | | |
| --- | --- | --- | --- | --- | --- | --- | --- |
|  |  |  | **France-like** | **Madagascar-like** | **France-like** | **Madagascar-like** | |
| $\beta_{h}$ | Human-to-human transmission rate | day^-1^ | $9\times{10}^{-3}$ | $11\times{10}^{-3}$ | *Calibrated* | | |
| $\gamma_{h}$ | Natural clearance rate | day^-1^ | $9.0\times{10}^{-3}$ | | (Haverkate et al., 2017)  *Supplementary note 8* | | |
| $\rho_{atb}$ | Antibiotic exposure rate | day^-1^ | $2.4\times{10}^{-3}$ | $3.3\times{10}^{-3}$ | (“Consommation d’antibiotiques en secteur de ville en France 2014-2024,” n.d.) *Supplementary note 6* | (Padget et al., 2017)  *Supplementary note 7* | |
| $\tau_{atb}$ | Average antibiotic treatment duration | day | $8$ | $4.1$ | *Fixed* | (Padget et al., 2017) | |
| $c$ | Antibiotic use effect on acquisition | / | $2.38$ | $2.2$ | (Tacconelli et al., 2020) | (Herindrainy et al., 2018) | |
| $\beta_{swim}$ | Transmission rate from swimming water to humans | day^-1^ | ${10}^{-4}$ | ${4\times10}^{-3}$ | *Fixed*  *Supplementary note 9* | *Fixed*  *Supplementary note 9* | |
| 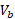 | Swallowed volume of water per swimming event | L | 0.027 | | (Schets et al., 2011) | | |
| $\beta_{veg}$ | Frequency of vegetable consumption | day^-1^ | ${0.5\times10}^{-4}$ | ${1\times10}^{-4}$ | *Fixed*  *Supplementary note 9* | | (*Profil des systèmes alimentaires - Madagascar*, 2021) *Supplementary note 9* |
| $\beta_{m}$ | Frequency of meat consumption | day^-1^ | ${0.5\times10}^{-4}$ | ${10}^{-5}$ | *Fixed*  *Supplementary note 9* | | (*Profil des systèmes alimentaires - Madagascar*, 2021) *Supplementary note 9* |
| $M$ | ESBL E.coli prevalence in meat | % | 17.8 | 57 | (Paradis et al., 2026) | (Milenkov et al., 2024) | |

### Table S2. Characteristics of parameters used for the vegetables model

| **Symbol** | **Parameter** | **Unit** | **Value** | | | | **References** | |
| --- | --- | --- | --- | --- | --- | --- | --- | --- |
|  |  |  | **France-like** | **Madagascar-like** | | | **France-like** | **Madagascar-like** |
| $\rho_{irr}$ | Irrigation exposition rate | day⁻¹ | 0.004 | | | 0.001 | *Fixed* | (“fao.org/aquastat/fr/countries-and-basins/country-profiles/country/MDG/,” n.d.) |
| $\tau_{irr}$ | Duration of irrigation | day | 120 | | | | *Fixed* | (“PDF 105 Filiere Fruits Et Legumes \| PDF \| Légumes \| Cuisine végan,” n.d.) |
| $h_{v}$ | Harvest rate of vegetables | day⁻¹ | $1/365$ | | | | *Fixed* | |
| $r_{V}$ | Plantation rate of vegetables | day⁻¹ | $1/365$ | | | | *Fixed* | |
| $\beta_{irr}$ | Transmission rate from surface water to vegetables | day⁻¹ | $0.04$ | | $0.06$ | | *Fixed* | *Fixed* |
| $\beta_{irr}^{tr}$ | Transmission rate treated water to vegetables | day⁻¹ | 0.016$\times\beta_{irr}$ | | $0.06$ | | (Mallard et al., 2025) | *Fixed* |
| $V_{irr}$ | Volume of water applied per irrigation event | L | ${10}^{-6}$ | | | | *Fixed* | |

### Table S3: Characteristics of parameters used for the water model

| **Symbol** | **Parameter** | **Unit** | **Value** | | **References** | |
| --- | --- | --- | --- | --- | --- | --- |
|  |  |  | **France-like** | **Madagascar-like** | **France-like** | **Madagascar-like** |
| $r_{w}$ | Growth rate of *E. coli* in water | day^-1^ | $0.2$ | | (Baker et al., 2016) | |
| $N_{max}$ | Carrying capacity in water | cells/L | 10^8^ | | (Sutradhar et al., 2021) | |
| $\beta_{w}$ | Gene transfer rate in water | day^-1^ | $1\times{10}^{-3}$ | | *Fixed* | |
| $\delta_{w}$ | Bacterial killing rate in response to antibiotics | day^-1^ | $0.02$ | | *Fixed* | |
| **Urban/hospital wastewater specific** | | | | | | |
| $\beta_{shed}$ | Rate of shedding into the sewage | day^-1^ | 10^3^ | | *Fixed* | |
| **Antibiotics in water** | | | | | | |
| 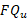  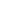 | Concentration of fluroquinolones in urban/ hospital wastewater | ng/L | 120 | 220 | (Wiest et al., 2018) | (Oharisi et al., 2023) |
| 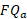 | Concentration of fluroquinolones in animal wastewater | ng/L | 0 | 0 | (Zahedi et al., 2021) | *No data, not in the common practices* |
| 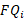 | Concentration of fluroquinolones in industrial wastewater | ng/L | 0 | 0 | *No industrial site* | |
| $E_{max}$ | Maximum effect of antibiotics on bacterial growth | / | 2 | | (Baker et al., 2016) | |
| $h$ | Hill coefficient | / | 2 | |  |  |
| $MIC_{s}$ | MIC for sensitive bacteria | $\mu g/L$ | 800 | |  |  |
| $MIC_{c}$ | MIC for resistant bacteria | $\mu g/L$ | 2000 | |  |  |
| **Treatment** | | | | | | |
| $effTP$ | Wastewater treatment efficiency | % | 90 | 20 | (“Observatoire des services publics de l’eau et de l’assainissement,” n.d.) | *Fixed* |
| **Wastewater flows** | | | | | | |
| $Q_{u}$ | Treated urban/hospital wastewater flow | m3/year | 14000000 | | Expert opinion | |
| $Q_{a}$ | Treated animal wastewater flow | m3/year | 60000 | | Expert opinion | |
| $Q_{i}$ | Treated industrial wastewater flow | m3/year | / | | No industrial site | |

### Table S4 Definition of model compartments and initial values in the different settings.

| **Symbol** | **Parameter** | **Unit** | **Value** | | **Reference setting** | |
| --- | --- | --- | --- | --- | --- | --- |
|  |  |  | **France-like** | **Madagascar-like** | **France-like** | **Madagascar-like** |
| $S_{u}$ | *E. coli* concentration in urban/hospital wastewater | CFU/100ml | 9e7 | 1e6 | Expert opinion | (Milenkov et al., 2024) |
| $C_{u}$ | ESBL *E. coli* concentration in urban/hospital wastewater | CFU/100ml | 8e6 | 1e5.06 | Expert opinion |  |
| $S_{a}$ | *E. coli* concentration in animal wastewater | CFU/100ml | 1e8 | 1e7.5 | (Dupouy et al., 2016) |  |
| $C_{a}$ | ESBL *E. coli* concentration in animal wastewater | CFU/100ml | 1e5 | 1e5.09 |  |  |
| $C_{d}$ | ESBL *E. coli* concentration in drinking water | CFU/100ml | 0 | 175 | *Fixed* | *Fixed* |
| $\frac{{C_{v}+S}_{v}}{N_{v}}$ | ESBL *E. coli* prevalence in vegetables | % | 6 | 27 | Expert opinion | (Rieux and Rabenandrasana, 2025) |
| $N_{h}$ | population |  | 121867 | 1600000 |  |  |
| $\frac{C+C^{A}}{N_{h}}$ | ESBL E.coli prevalence | % | 6 | 34 | (Bezabih et al., 2021; Woerther et al., 2013) | (Milenkov et al., 2024) |
| 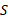 | Non-carriers |  | 112549 | 1050545 | *Supplementary note 5* | |
| 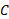 | Carriers |  | 6978 | 527807 |  |  |
| 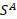 | Non-carriers under antibiotic treatment |  | 2006 | 5455 |  |  |
| 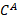 | Carriers under antibiotic treatment |  | 334 | 16193 |  |  |

### Supplementary Note 1. Equations of the ordinary differential equation system for the general population model.

$$\frac{dS_{h}}{dt}=-\beta_{h}S_{h}\frac{C_{h}^{A}+C_{h}}{N_{h}}-\lambda_{env}S_{h}+\gamma_{h}C_{h}-\rho_{atb}S_{h}+\frac{1}{\tau_{atb}}S_{h}^{A}$$

$$\frac{dC_{h}}{dt}=\beta_{h}S_{h}\frac{C_{h}^{A}+C_{h}}{N_{h}}+\lambda_{env}S_{h}-\gamma_{h}C_{h}-\rho_{atb}C_{h}+\frac{1}{\tau_{atb}}C_{h}^{A}$$

$$\frac{dS_{h}^{A}}{dt}=-{c\beta}_{h}S_{h}^{A}\frac{C_{h}^{A}+C_{h}}{N_{h}}-{c\lambda}_{env}S_{h}^{A}+\gamma_{h}C_{h}^{A}+\rho_{atb}S_{h}-\frac{1}{\tau_{atb}}S_{h}^{A}$$

$$\frac{dC_{h}^{A}}{dt}={c\beta}_{h}S_{h}^{A}\frac{C_{h}^{A}+C_{h}}{N_{h}}+{c\lambda}_{env}S_{h}^{A}-\gamma_{h}C_{h}^{A}+\rho_{atb}C_{h}-\frac{1}{\tau_{atb}}C_{h}^{A}$$

**Forces of infections:**

$$\lambda_{env}=\lambda_{swim}+ \lambda_{drinking}+ \lambda_{veg}{+\lambda}_{meat}$$

*Acquisition of ESBL-Ec through bathing activities*

$$\lambda_{swim}=\beta_{swim}\left( 1-exp \left( -C_{s}.V_{s} \right) \right)$$

where$\beta_{swim}$ represents the transmission rate from contaminated water to human non-carriers. In other words, $\beta_{swim}$ corresponds to the individual bathing rate per day multiplied by the risk of ESBL-Ec acquisition following ingestion of contaminated water. $C_{s}$ corresponds to the ESBL-Ec concentration (CFU/100mL) in surface water and $V_{s}$ to the swallowed volume of water per person per swimming event which are together formulated as the exposure probability $\left( 1-exp \left( -C_{s}*V_{s} \right) \right)$ (Schijven et al., 2015).

*Acquisition of ESBL-Ec via drinking water.* The human exposure to ESBL-Ec via drinking water consumption was modelled with the force of acquisition written as:

$$\lambda_{drinking}=\beta_{drink}\left( 1-exp \left( -C_{d}.V_{d} \right) \right)$$

where$\beta_{drink}$ represents the transmission rate from colonized water to human non-carriers, defined as the product of the drinking rate per individual per day and the probability of ESBL-Ec acquisition following ingestion of contaminated water. $C_{d}$ represents the ESBL-Ec concentration in drinking water and $V_{d}$to the volume of water per person per drinking event.

*Acquisition of ESBL-Ec via vegetable consumption****.*** Rising temperatures are anticipated to reduce groundwater availability and increase the reuse of treated wastewater for irrigation, thereby influencing the carriage of ESBL-Ec on vegetables entering the food chain. Exposure through vegetable consumption was modeled as:

$$\lambda_{veg}=\beta_{veg}\frac{C_{v}^{I}+C_{v}}{N_{v}}$$

Where $\beta_{veg}$ corresponds to the transmission rate between vegetables and humans. It is defined as the daily frequency of vegetable consumption per individual multiplied by the probability of acquiring the resistant strain following consumption of contaminated vegetables. $\frac{C_{v}^{I}+C_{v}}{N_{v}}$ is the prevalence of ESBL-Ec in vegetables at time t.

*Acquisition of ESBL-Ec via consumption of animal products.* The acquisition of ESBL-Ec via animal products (meat) was modeled as:

$$\lambda_{meat}=\beta_{meat}C_{m}$$

Where $\beta_{meat}$ corresponds to the transmission rate from contaminated meat to human non-carriers. It is defined as the daily frequency of meat consumption per individual multiplied by the probability of acquiring the resistant strain following the ingestion of contaminated meat, while $C_{m}$ represents the prevalence of contamination in meat. Meat contamination ($C_{m}$) was considered an external input rather than a dynamic model component, as the present model focuses on the human–environment interface and does not explicitly represent animal reservoirs or food-production processes.

### Supplementary Note 2. Equations of the ordinary differential equation system for the vegetables model.

$\frac{\mathrm{dS}_{v}}{dt}=r_{v} (S_{v}+C_{v})+\frac{1}{\tau_{irr}}S_{v}^{I}-\rho_{irr}S_{v} -\gamma_{v}S_{v}$

$$\frac{dS_{v}^{I}}{dt}=\rho_{irr}S_{v}-\lambda_{irr}S_{v}^{I}- \frac{1}{\tau_{irr}}S_{v}^{I}$$

$$\frac{dC_{v}^{I}}{dt}=\lambda_{irr}S_{v}^{I}- \frac{1}{\tau_{irr}}C_{v}^{I}$$

$$\frac{dC_{v}}{dt}=\frac{1}{\tau_{irr}}C_{v}^{I}-\gamma_{v}C_{v}$$

**Force of infection:**

The probability of vegetable contamination with ESBL-Ec during irrigation was modelled using a dose-response approach adapted from (Schijven et al., 2015), analogous to the human exposure model. Vegetables were assumed to be irrigated with surface water, treated urban wastewater and treated animal wastewater. The corresponding transmission rates are denoted by$\beta_{irr}^{s}$for surface water and $\beta_{irr}^{tr}$ for treated wastewater. $\beta_{irr}^{tr}$ and $\beta_{irr}^{s}$ both are rates defined as the product of the irrigation rate per day and the probability of vegetable contamination following an irrigation event. $V_{irr}$ represents the volume of water applied during a single irrigation event. The force of contamination through irrigation, defined as $\lambda_{irr}$, is given by:

$$\lambda_{irr}=\beta_{irr}^{tr}\left( 1-exp \left( -C_{u}^{tr}.V_{irr} \right) \right)+\beta_{irr}^{tr}\left( 1-exp \left( -C_{a}^{tr}.V_{irr} \right) \right)+ \beta_{irr}^{s}\left( 1-exp \left( -C_{s}.V_{irr} \right) \right)$$

While $\left( 1-exp \left( -C_{u}^{tr}.V_{irr} \right) \right)$ represent the probability of exposure.

Equations of the ordinary differential equation system for the vegetables model are present in Supplementary Note 2.

### Supplementary Note 3. Equations of the ordinary differential equation system for the wastewater model.

$$\frac{dS_{w}}{dt}=r_{w}\left( 1-\frac{S_{w}+C_{w}}{N_{max}} \right)e_{s}{(t)S}_{w}-\beta_{w}S_{w}\frac{C_{w}}{S_{w}+C_{w}}+ \gamma_{w} C_{w}-\delta_{w}S_{w}-\lambda_{h,w}S_{w}$$

$$\frac{dC_{w}}{dt}=r_{w}\left( 1-\frac{S_{w}+C_{w}}{N_{max}} \right)e_{r}{(t)C}_{w}+\beta_{w}S_{w}\frac{C_{w}}{S_{w}+C_{w}}- \gamma_{w} C_{w}-\delta_{w}C_{w}+\lambda_{h,w}S_{w}$$

With:

$$e_{s}(t)= 1-\frac{E_{max}FQ_{w}(t)}{FQ_{w}(t)+ MIC_{s}}$$

$$e_{r}(t)= 1-\frac{E_{max}FQ_{w}(t)}{FQ_{w}(t)+ MIC_{r}}$$

where $FQ_{w}$ denotes the fluoroquinolone concentration in water and $MIC_{s}$ and $MIC_{r}$ represent the minimum inhibitory concentrations of susceptible and resistant bacteria, respectively and $E_{max}$is the maximum antibiotic effect.

**Forces of infection:**

Human shedding contributes through feces to the contamination of the urban and hospital ($u)$ wastewater compartment with ESBL-Ec. This process is represented by the force of human-to-water contamination, $\lambda_{h}$, defined as:

$$\lambda_{h,u}=\beta_{shed}\frac{C_{h}^{A}+ C_{h}}{N_{h}}$$

Here $\frac{C_{h}^{A}+ C_{h}}{N_{h}}$ denotes the prevalence in the human population colonized with ESBL-Ec over time, and $\beta_{shed}$represents the rate at which resistant bacteria are released into the wastewater system through human shedding.

The human shedding affects the urban/hospital ($u)$ wastewater compartment but does not directly contribute to animal wastewater ($a)$, industrial wastewater ($i)$ in the present model.

$$\boldsymbol{\lambda}_{\mathbf{h}\mathbf{,}\mathbf{a}}\mathbf{=}\boldsymbol{\lambda}_{\mathbf{h}\mathbf{,}\mathbf{i}}\mathbf{=}\mathbf{0}$$

### Supplementary Note 4. Wastewater treatment, mixing, and bacterial decay in surface waters

**Wastewater treatment. Mixing of treated effluents.**

The treated effluents are subsequently discharged into the receiving water body, where they mix according to their relative flow rates. The total wastewater flow is given by:

$$Q_{tot}=Q_{u}+Q_{a}+Q_{i}$$

where $Q_{u}$, $Q_{a}$ and $Q_{i}$ denote the treated urban/hospital, animal, and industrial wastewater flows, respectively. The concentration immediately after mixing is calculated as the flow-weighted average:

$$C_{mix}=\frac{Q_{u}C_{u}^{tr}+Q_{a}C_{a}^{tr}+Q_{i}C_{i}^{tr}}{Q_{tot}}$$

where $C_{mix}$ is the ESBL-Ec concentration in the receiving water immediately downstream of the discharge point.

**Environmental decay**

Following discharge, bacterial concentrations in the receiving water decrease over time through environmental decay, as described by (Schijven et al., 2015). The concentration at the potential human exposure site, denoted ($C_{s})$, is computed as :

$$C_{s}\left( t \right)=C_{mix}exp\left( -{10}^{-1.04+0.017T+0.5}t \right) D$$

Where $C_{s}$ is the ESBL-Ec concentration in surface water after a travel time ($t$), $T$ is the water temperature, $t$ is the travel time in days and $D$is a dilution factor. A travel time of 10 days was assumed between the WWTP discharge and the potential site of human exposure and a dilution factor $D$ of up to 10^9^ time (Schijven et al., 2015).

The concentration remaining after treatment if therefore :

$$C_{w}^{tr}=(1-{eff}_{TP}) C_{w}$$

where $C_{w}^{tr}$ is the ESBL-Ec concentration in the treated effluent.

### Supplementary Note 5. Human Model Initialization

The human population is stratified into four compartments according to colonization status and antibiotic exposure: non-colonized without antibiotics ($S^{0}$), colonized without antibiotics ($C^{0}$), non-colonized under antibiotic treatment ($S^{A}$), and colonized under antibiotic treatment ($C^{A}$). The total population size is fixed as $N$.

$$N=C^{A}+C+S^{A}+S$$

Baseline ESBL-Ec colonization prevalence was assumed to be $p$:

$$C^{A}+C=p*N$$

So that means 7312 people in France and 544000 people in Madagascar.

The total number of individuals under treatment is:

$$S^{A}+C^{A}=\rho_{\mathrm{atb}}\times\tau_{\mathrm{atb}}\times N=N_{A}.$$

So that means 2340 people in France and 21648 in Madagascar.

Antibiotic consumption increases the risk of ESBL-Ec acquisition by a factor *c*.

Thus $C^{A}=c\times p\times N_{A}$.

That means 334 people in France and 16193 people in Madagascar.

### Supplementary Note 6. Antibiotic exposure rate computation in France

We consider the proportion of individuals $\rho_{\mathrm{atb}}$who initiate antibiotic therapy each day, with treatment lasting for a fixed duration of $\tau_{\mathrm{atb}}$days. Using the annual rate of systemic antibiotic prescriptions in Doubs in 2024 (865.011 prescriptions per 1000 inhabitants per year), the daily initiation rate was estimated as

$$\rho_{\mathrm{atb}}=\frac{865.011}{1000\times365}=2.4\times{10}^{-3} \text{day}^{-1}$$

We assumed a fixed treatment duration of $\tau_{\mathrm{atb}}=8$days.

### Supplementary Note 7. Antibiotic exposure rate computation in Madagascar

Based on a community survey of antibiotic consumption among children in Madagascar and Senegal (Padget et al., 2017), the estimated proportion of the population in Antananarivo who had consumed antibiotics during the previous three months (90 days) was 29.3% (95% CI: 25.0–34.1%). The corresponding daily antibiotic treatment initiation rate was calculated as:

$$\rho_{\mathrm{atb}}=\frac{0.293}{90}= 3.3\times{10}^{-3} \text{day}^{-1}$$

The survey found a length of antibiotic consumption of $\tau_{\mathrm{atb}}=$4.1 days.

### Supplementary Note 8. Human decolonization rate

For both settings, under the baseline scenario, the median carriage duration was taken from a study in the community that looked at discharged patients within households in the Netherlands $(Haverkate et al., 2017)$. The median carriage duration for ESBL-producing Enterobacterales for household members was of 111 days (56-217).

The decolonization rate was thus defined as :

$$\gamma_{h}=\frac{1}{111}=9.0\times{10}^{-3} \text{day}^{-1}$$

### Supplementary Note 9. Environment-to-human transmission rates

(Haverkate et al., 2017) estimated a background environmental acquisition rate of 1.5 × 10⁻⁴ per day. In the present framework, environmental exposure is considered to occur through three main pathways: swimming in surface freshwater, consumption of drinking water, and consumption of raw vegetables. Hereafter, $F$and $M$refer to France and Madagascar, respectively.

(Mughini-Gras et al., 2019) estimated source attribution for intestinal carriage of ESBL-producing *Escherichia coli* in the community in the Netherlands. Their results suggested that raw vegetable consumption accounted for approximately 1.1% of cases, while exposure related to swimming in surface freshwater accounted for approximately 2–3%. Based on these estimates, we assumed that the transmission rate from swimming event was higher than raw vegetable consumption:

$\beta_{swim}>\beta_{veg}$.

In coherence with (Haverkate et al., 2017) we fixed $\beta_{veg}(F)= 0.5 \times10⁻⁴$ day^-1^ and $\beta_{swim}(F)= 1$ × 10⁻⁴day^-1^ in the French setting.

Setting-specific assumptions

**Vegetables consumption.** In Madagascar, diets are mainly based on rice, cassava, maize and sweet potato; rice accounts for around 70% of cereal production. (*Profil des systèmes alimentaires - Madagascar*, 2021). Vegetable consumption is therefore generally more central than in France, where it was estimated at 140.6 g/person/day in 2024.

$$\beta_{veg}\left( M \right)>\beta_{veg}(F)$$

**Meat consumption.** In terms of meat consumption it’s 38g/pers/day in Madagascar (*Profil des systèmes alimentaires - Madagascar*, 2021). Whereas is we eat meat 1.5 times per day.

$$\beta_{meat}\left( F \right)>\beta_{meat}(M)$$

**Bathing activities.**

$\beta_{swim}$ corresponds to the bathing rate per individual per day times the risk of acquisition following the ingestion of contaminated water. The risk of acquisition following the ingestion of contaminated water is similar in the two settings. However, the bathing rate is different. According to (Schets et al., 2011), freshwater and seawater sites were visited 6 to 8 times a year for one person in the Netherlands. In contrast, evidence from Madagascar indicates much more frequent freshwater contact. In Marolambo District, eastern Madagascar, 40% of schoolchildren reported contact with the Nosivolo River daily or more frequently in 2017, increasing to 80% in 2018 (Spencer et al., 2022). The study also reports that children use the river and surrounding streams for washing, bathing, drinking, water collection, farming, and other activities. For the illustrative Madagascar setting, we assumed 0.8 freshwater contacts per day (approximately 292 contacts per year). Thus the contact rate is 42 times higher for Madagascar.

$\beta_{swim}(M)= 4$ × 10⁻^3^day^-1^

### Table S5. Antibiotic scenarios results in France

Percentage change relative to the baseline scenario.

| **scenario** | **Human_change_pct** | **Vegetable_change_pct** | **C_u_change_pct** |
| --- | --- | --- | --- |
| Rising human use | 48.90 | 14.86 | 30.50 |
| Environmental contamination | 29.71 | 56.91 | 227.12 |
| Coupled increase | 73.27 | 58.28 | 246.10 |

### Table S6. Antibiotic scenarios results in Madagascar

Percentage change relative to the baseline scenario.

| **scenario** | **Human_change_pct** | **Vegetable_change_pct** | **C_u_change_pct** |
| --- | --- | --- | --- |
| Rising human use | 3.94 | 0.04 | 1.84 |
| Environmental contamination | 16.11 | 0.91 | 106.23 |
| Coupled increase | 21.58 | 0.94 | 129.36 |

### Table S7. Climate change scenarios in France

Percentage change relative to the baseline scenario.

| **scenario** | **Human_change_pct** | **Vegetable_change_pct** | **C_u_change_pct** |
| --- | --- | --- | --- |
| Climate change | 2.68 | 8.25 | 14.48 |
| Climate change+ Flood | 23.18 | 50.80 | 31.22 |
| Climate change+ Water reuse | 3.90 | 32.90 | 15.51 |
| Climate change+ Drought | 13.26 | 21.92 | 45.37 |
| All | 40.55 | 91.76 | 69.47 |

### Table S8. Climate change scenarios in Madagascar

Percentage change relative to the baseline scenario.

| **scenario** | **Human_change_pct** | **Vegetable_change_pct** | **C_u_change_pct** |
| --- | --- | --- | --- |
| Climate change | 1.89 | 0.53 | 9.08 |
| Climate change+ Flood | 7.47 | 1.12 | 12.17 |
| Climate change+ Water reuse | 1.90 | 2.46 | 9.09 |
| Climate change+ Drought | 11.72 | 0.80 | 27.87 |
| All | 17.82 | 3.07 | 31.16 |

PDF 105 Filiere Fruits Et Legumes | PDF | Légumes | Cuisine végan [WWW Document], n.d. . Scribd. URL https://fr.scribd.com/document/1028583436/PDF-105-Filiere-Fruits-Et-Legumes (accessed 8.7.26).

Profil des systèmes alimentaires - Madagascar, 2021. . FAO; European Union ; CIRAD; https://doi.org/10.4060/cb6861fr

Rieux, A., Rabenandrasana, M.A.N., 2025. Identification of ESBL ‐Producing Enterobacterales From Vegetable Plants: Preliminary Findings From a Small Cross‐Sectional Study in a Rural Area of Madagascar. Environ Microbiol Rep 17, e70130. https://doi.org/10.1111/1758-2229.70130

Schets, F.M., Schijven, J.F., de Roda Husman, A.M., 2011. Exposure assessment for swimmers in bathing waters and swimming pools. Water Research 45, 2392–2400. https://doi.org/10.1016/j.watres.2011.01.025

Schijven, J.F., Blaak, H., Schets, F.M., de Roda Husman, A.M., 2015. Fate of Extended-Spectrum β-Lactamase-Producing Escherichia coli from Faecal Sources in Surface Water and Probability of Human Exposure through Swimming. Environ. Sci. Technol. 49, 11825–11833. https://doi.org/10.1021/acs.est.5b01888

Spencer, S.A., Andriamasy, E.H., Linder, C., Penney, J.M.S., Henstridge-Blows, J., Russell, H.J., Hyde, K., Sheehy, C., Young, I.L., Sjoflot, B., Rakotomampianina, D.A.L., Nandimbiniaina, A.M., Raderalazasoa, G.U., Ranaivoson, T.N., Andrianiaina, A., Michèle, R.S.M., Rohe, Z.A., Bustinduy, A.L., Stothard, J.R., Cruickshank, S.M., Edosoa, G.T., Rahetilahy, A.M., 2022. Impact of a Novel, Low-Cost and Sustainable Health Education Program on the Knowledge, Attitudes, and Practices Related to Intestinal Schistosomiasis in School Children in a Hard-to-Reach District of Madagascar. The American Journal of Tropical Medicine and Hygiene 106, 685–694. https://doi.org/10.4269/ajtmh.21-0220

Sutradhar, I., Ching, C., Desai, D., Suprenant, M., Briars, E., Heins, Z., Khalil, A.S., Zaman, M.H., 2021. Computational Model To Quantify the Growth of Antibiotic-Resistant Bacteria in Wastewater. mSystems 6, 10.1128/msystems.00360-21. https://doi.org/10.1128/msystems.00360-21

Tacconelli, E., Górska, A., De Angelis, G., Lammens, C., Restuccia, G., Schrenzel, J., Huson, D.H., Carević, B., Preoţescu, L., Carmeli, Y., Kazma, M., Spanu, T., Carrara, E., Malhotra-Kumar, S., Gladstone, B.P., 2020. Estimating the association between antibiotic exposure and colonization with extended-spectrum β-lactamase-producing Gram-negative bacteria using machine learning methods: a multicentre, prospective cohort study. Clinical Microbiology and Infection 26, 87–94. https://doi.org/10.1016/j.cmi.2019.05.013

Wiest, L., Chonova, T., Bergé, A., Baudot, R., Bessueille-Barbier, F., Ayouni-Derouiche, L., Vulliet, E., 2018. Two-year survey of specific hospital wastewater treatment and its impact on pharmaceutical discharges. Environ Sci Pollut Res 25, 9207–9218. https://doi.org/10.1007/s11356-017-9662-5

Woerther, P.-L., Burdet, C., Chachaty, E., Andremont, A., 2013. Trends in Human Fecal Carriage of Extended-Spectrum β-Lactamases in the Community: Toward the Globalization of CTX-M. Clinical Microbiology Reviews 26, 744–758. https://doi.org/10.1128/cmr.00023-13

Zahedi, S., Gros, M., Balcazar, J.L., Petrovic, M., Pijuan, M., 2021. Assessing the occurrence of pharmaceuticals and antibiotic resistance genes during the anaerobic treatment of slaughterhouse wastewater at different temperatures. Science of The Total Environment 789, 147910. https://doi.org/10.1016/j.scitotenv.2021.147910
